# Regional disparities in prescription methamphetamine and amphetamine distribution across the United States in 2019

**DOI:** 10.1101/2022.05.23.22275471

**Authors:** Sarah D. Lopera, Victoria M. O’Kane, Jessica L. Goldhirsh, Brian J. Piper

## Abstract

**Introduction:** Methamphetamine is a highly addictive psychostimulant and controlled substance that has detrimental health consequences for chronic users. Amphetamine is a structurally-related stimulant commonly used to treat ADHD, however, it also has a high risk for substance misuse. The objectives of this report were to characterize the regional differences in prescription methamphetamine and amphetamine distribution in the US, and examine potential reasons for variations in distribution.

**Methods:** Data for prescription methamphetamine and amphetamine distribution was obtained from the US Drug Enforcement Administration’s Automation of Reports and Consolidated Orders System (ARCOS) Retail Drug Summary Report for 2019. Quarterly, state, and regional differences in distributions of the two controlled substances were analyzed and.

**Results:** The preponderance (97%) of retail drug distribution for both drugs in 2019 were made through pharmacies. In the same year, prescription methamphetamine (+6.8%) and amphetamine (+5.8%) saw increases in drug distribution from Quarter 1 to Quarter 4.. Across the entire US, total per capita drug weight distribution of amphetamine was exactly 4,000 times higher than methamphetamine. Regionally, total per capita drug weight for methamphetamine was highest in the West (32.2% of total distribution) and lowest in the Northeast (17.4%). The total per capita drug weight for amphetamine was highest in the South (37.0% of total distribution) and lowest in the Northeast (19.4%). The ratio between the 90th and 10th percentiles of per capita drug weight by state was 4.39 for methamphetamine and 2.45 for amphetamine. Distribution of methamphetamine only measured 16.1% of its production quota, while distribution of amphetamine measured 54.0% of its production quota.

**Discussion:** Overall, prescription amphetamine distribution was common while prescription methamphetamine distribution was rare. Regional disparities were also present with lowest distribution of both substances in the Northeast region. The patterns observed in distribution are likely the result of stigmatization, differences in accessibility, and the efforts of initiatives such as the Montana Meth Project. This cycle will likely also change in response to physician recommendation and public opinions surrounding issues for pharmacotherapies for ADHD management.

## 1. Introduction

Methamphetamine, a potent amphetamine derivative, is a well-recognized psychostimulant that acts upon the central nervous system inducing feelings of euphoria, increased alertness, mood and concentration, as well as elevated interest in external stimuli [1]. This highly addictive (Schedule II) drug has a higher lipid solubility, making it capable of crossing the blood brain barrier rapidly, and produces a prolonged action due to greater elimination half-life, on the order of 10-20 hours, when compared to cocaine, which is measured in minutes to an hour, and other amphetamines (11-14 hours for d-AMP and l-AMP) [2-4]. Once introduced into circulation, concentration of the drug rapidly accumulates within the brain and leads to the release of various monoamines, primarily, dopamine, serotonin, and noradrenaline, which in turn affect neurochemical processes responsible for reward processing, motivation and attention, ultimately promoting drug dependence [5,6]. Methamphetamine is available in many different forms, such as in crystalline hydrochloride salt form, referred to as crystal meth, or as a prescription drug in tablet form [5]. Extra-medical use of the drug is typically in crystalline form and occurs in significantly higher doses than what is prescribed orally as a controlled medical treatment. Faster onset of the drug is typically achieved via alternative methods of administration, such as intravenous, intranasal, vaginal, and inhalation, in which users consume higher doses for recreational use prompted by socio-cultural influences or for performance enhancement [6].

Neurotoxicity of methamphetamine exposure has been supported by abnormalities in brain structure and physiological manifestations, such as hallucinations, delusions, depression, suicidality and aggression [6]. Furthermore, addiction has been known to occur faster than with cocaine and to a stronger extent in which a single exposure is enough to establish dependency [6]. Unlike other stimulant drugs, drug-seeking behaviors for methamphetamine persist even after tolerance has been reached [6]. Physiological effects have been reported to include neuronal tissue death and damage, dental disease, cardiovascular morbidity, stroke, increased prevalence of STIs, such as HIV infection, and cognitive impairment [6]. Other neurological manifestations of heavy meth use includes fromications, in which there is a sensation of insects crawling on or under the skin, which may lead to skin-picking behaviors that form lesions and ulcers. Lack of hygiene combined with consistent disruption of wound-healing mechanisms has resulted in a high incidence of cellulitis and skin infection in those who misuse the drug. Furthermore, heavy methamphetamine use disrupts the immunological system, altering host immunity by increasing the pH of acidic organelles, which inhibits phagocytosis and antigen presenting processes essential in innate immunity, decreasing one’s ability to fight off infection [3]. Overall, long-term and chronic methamphetamine use has shown detrimental psychosocial and physiological consequences making it a major public health concern.

Despite the addictive nature of the drug and its implications on public health, therapeutic uses of amphetamine due to its ability to increase alertness and concentration in order to treat adult patients with attention-deficit hyperactivity disorder (ADHD) and narcolepsy. Compared to non-stimulant drugs such as clonidine and atomoxetine, stimulant drugs such as amphetamine-based agents (Adderall, Ritalin, Dexedrine) are more effective in treating adult-ADHD by increasing catecholamine availability in striatal and cortical brain regions [7]. The fine line between drug misuse and drug therapy can be noted by the prevalence of adults with ADHD who also suffer from methamphetamine misuse, even when amphetamine is being used as an effective treatment option. Substance misuse disorders have been noted to co-occur with ADHD and subsequent substance misuse has also been noted in which methamphetamine misuse replaces ADHD medication dependency [8,9]. The major issue within this complicated relationship between the medical use of amphetamine and stimulant misuse disorders involving methamphetamine is that it has led to the stigmatization of both methamphetamine and amphetamine and consequently, the underreporting of use. Prior pharmacoepidemiological research has examined changes and regional disparities in amphetamine, but less is known for methamphetamine [10,14,21]. In this study, we aimed to demonstrate the variable weight distribution of methamphetamine and amphetamine across the United States, and provide a regional analysis.

## 2. Methods

### 2.1 Procedures

The Drug Enforcement Administration’s (DEA) Automation of Reports and Consolidated Orders System (ARCOS) was used to obtain drug weight data for methamphetamine and amphetamine by US state for the year 2019, as well as the production quotas for the same year. This year was chosen as the most recent that this information was reported for both substances. Methamphetamine and amphetamine are both Schedule II controlled substances, which means transactions from manufactures through the drug supply chain to point-of-sale distributors are required by law to be reported to ARCOS. Due to their structural similarity, amphetamine data was included for comparison to methamphetamine. ARCOS had also shown high-concordance with a state prescription drug monitoring program for ADHD medication previously [10]. Our data was obtained specifically from Reports 3, 4, and 7 of the 2019 Retail Drug Summary Report. This information is publicly available at [11]. Data regarding production quotas for 2019 was obtained from the DEA’s notice on the Established Aggregate Production Quotas for Schedule I and II Controlled Substances… for 2019, which is also publicly available at [12]. Procedures were approved as exempt by the IRB at Geisinger.

### 2.2 Data Analysis

The total per capita quarterly drug distributions, measured as (mg) per 1,000 people were calculated for both methamphetamine and amphetamine. The 50 US states were divided into the following four regions based on the grouping used by the US Census Bureau: Northeast, South (including Washington DC), Midwest, and West [13]. The total per capita drug weights for each state and region, measured as (mg) per 1,000 people, were calculated for both methamphetamine and amphetamine. Heat maps were generated with http://www.heatmapper.ca/geomap/ using the total per capita drug weights measured as (mg) per 1,000 people. Per capita distribution of both substances was also expressed as a ratio relative to the lowest, nonzero, state. A 90/10 percentile ratio for each substance was also determined. Percentage data from retail drug purchases for methamphetamine and amphetamine, separated by business activity, was determined. The ratio of substance distributed in 2019 relative to production quotas for 2019 was determined for both drugs. Figures were prepared with GraphPad Prism, 9.3.1.

## 3. Results

In Figure 1, the total per capita quarterly drug distributions of both methamphetamine (+6.8%) and amphetamine (+5.8%) increased from Q1 to Q4. However, the increase was more consistent from quarter to quarter with amphetamine, but not so with methamphetamine. The total per capita drug distribution of methamphetamine in Q2 saw a decrease compared to Q1 (-5.1%), but was followed by increases from Q2 to Q3 (+11.3%) and Q3 to Q4 (+1.1%). The ratio of amphetamine to methamphetamine distribution was 3,976.9 in Q1, 4,298.6 in Q2, 3,908.8 in Q3, and 3,939.7 in Q4 (Mean = 4,031.0).

**Figure 1.**
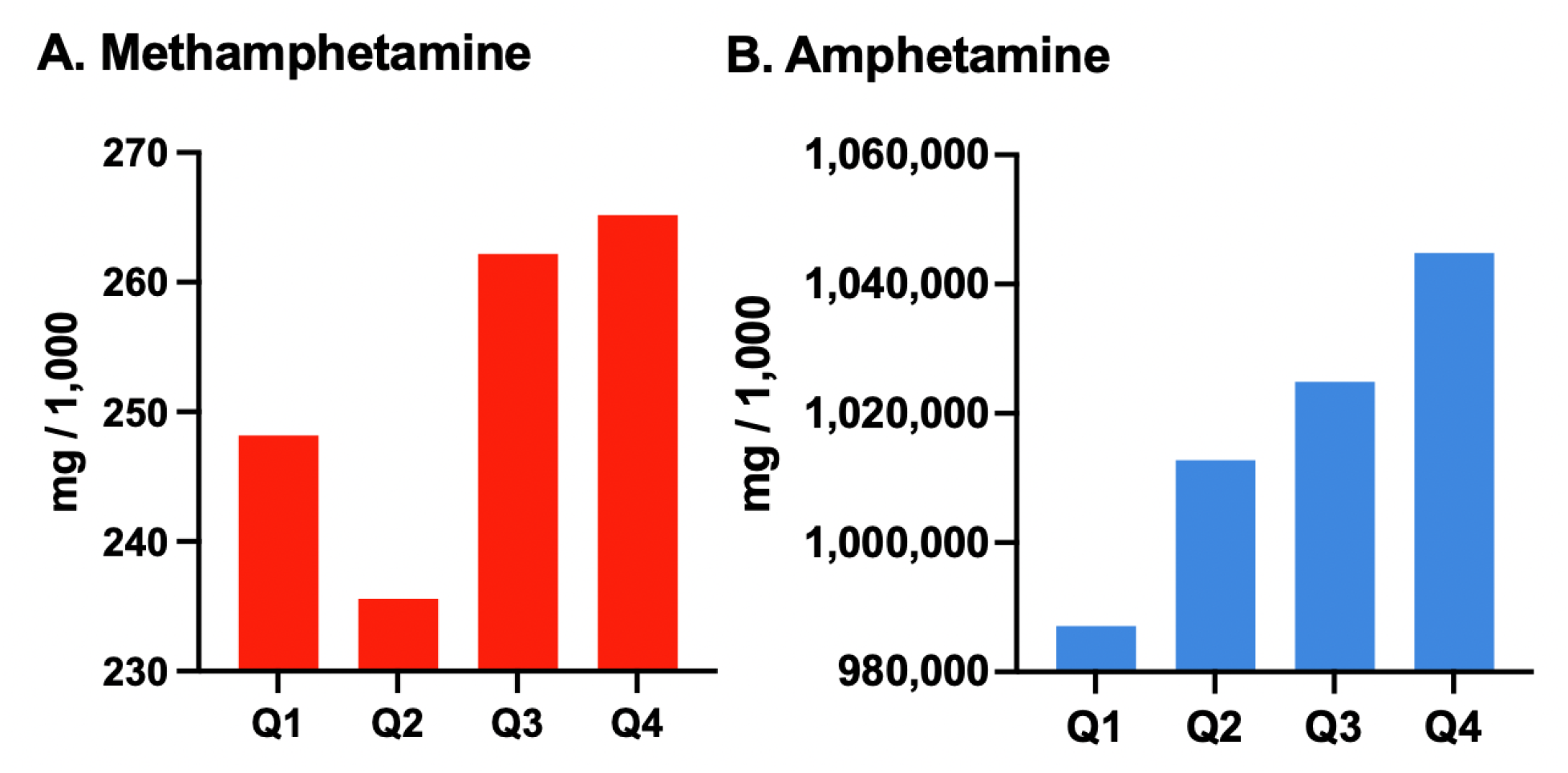
Total per capita quarterly drug distribution of (A) methamphetamine and (B) amphetamine in milligrams per 1,000 people in the US in 2019 as reported to the Drug Enforcement Administration’s Automated Reports and Consolidated Orders System. Note restricted Y-axis range to improve viewability.

Figure 2 shows the geographical distribution of each agent. It can be noted that for the state of Montana distribution of prescription methamphetamine was zero, but distribution of amphetamine was nonzero. The top (highest) and bottom (lowest) three states for each drug can be noted in Figure 2, but more easily identified in Table 1, which expresses the per capita distributions as a ratio relative to the lowest, nonzero, state. Comparing the state with the highest distribution of prescription methamphetamine to the state with the lowest, Washington had 16.8 times more per capita distribution than Arkansas. For amphetamine, the range was not nearly as widespread, with Louisiana having only 4.92 times more per capita distribution than New Mexico. When the ratio between the 90th and 10th percentiles was determined for both substances, the following was observed. For methamphetamine, states with the top 90% of prescription methamphetamine distribution had 4.39-fold more of the drug than the states with the bottom 10%. For amphetamine, states with the top 90% had 2.45-fold more drug than the bottom 10%.

**Figure 2.**
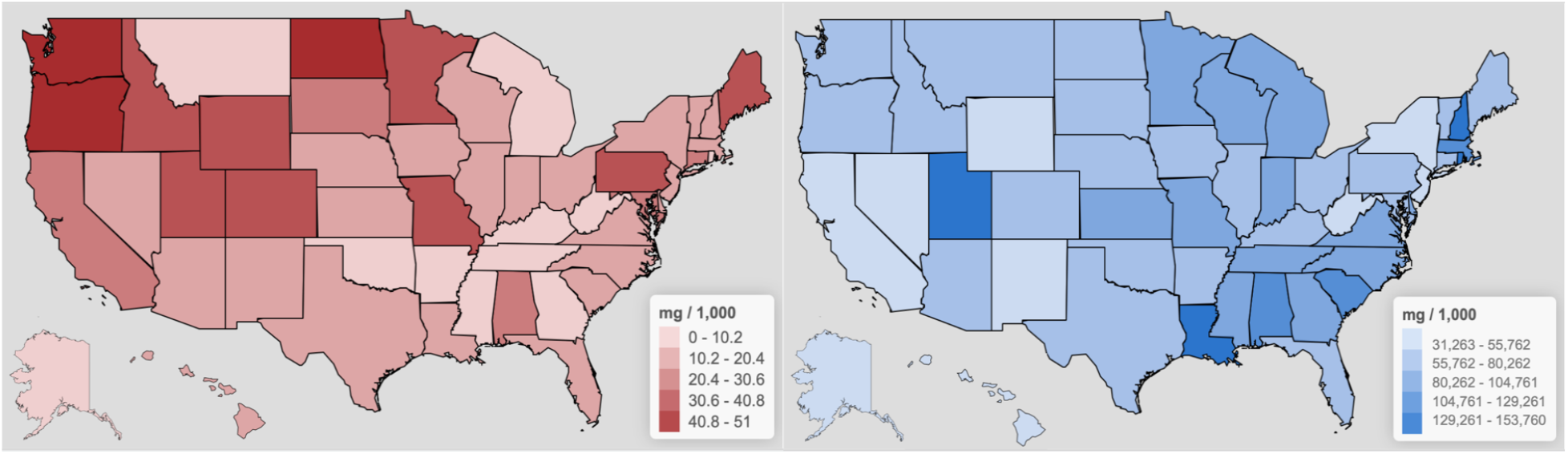
Heat maps of the per capita drug weights of methamphetamine (left) and amphetamine (right) in milligrams per 1,000 people by US state in 2019.

In Figure 3, the total per capita drug weight of methamphetamine was greatest in the West, followed by the Midwest, then the South, and finally the Northeast. The total per capita drug weight of methamphetamine in the West was nearly twice as high as that in the Northeast. The total per capita drug weight of amphetamine was greatest in the South, followed by the Midwest, then the West, and finally the Northeast. Again, the total per capita drug weight of amphetamine in the lowest region, the South, was nearly twice as high as that in the highest region, the Northeast.

**Figure 3.**
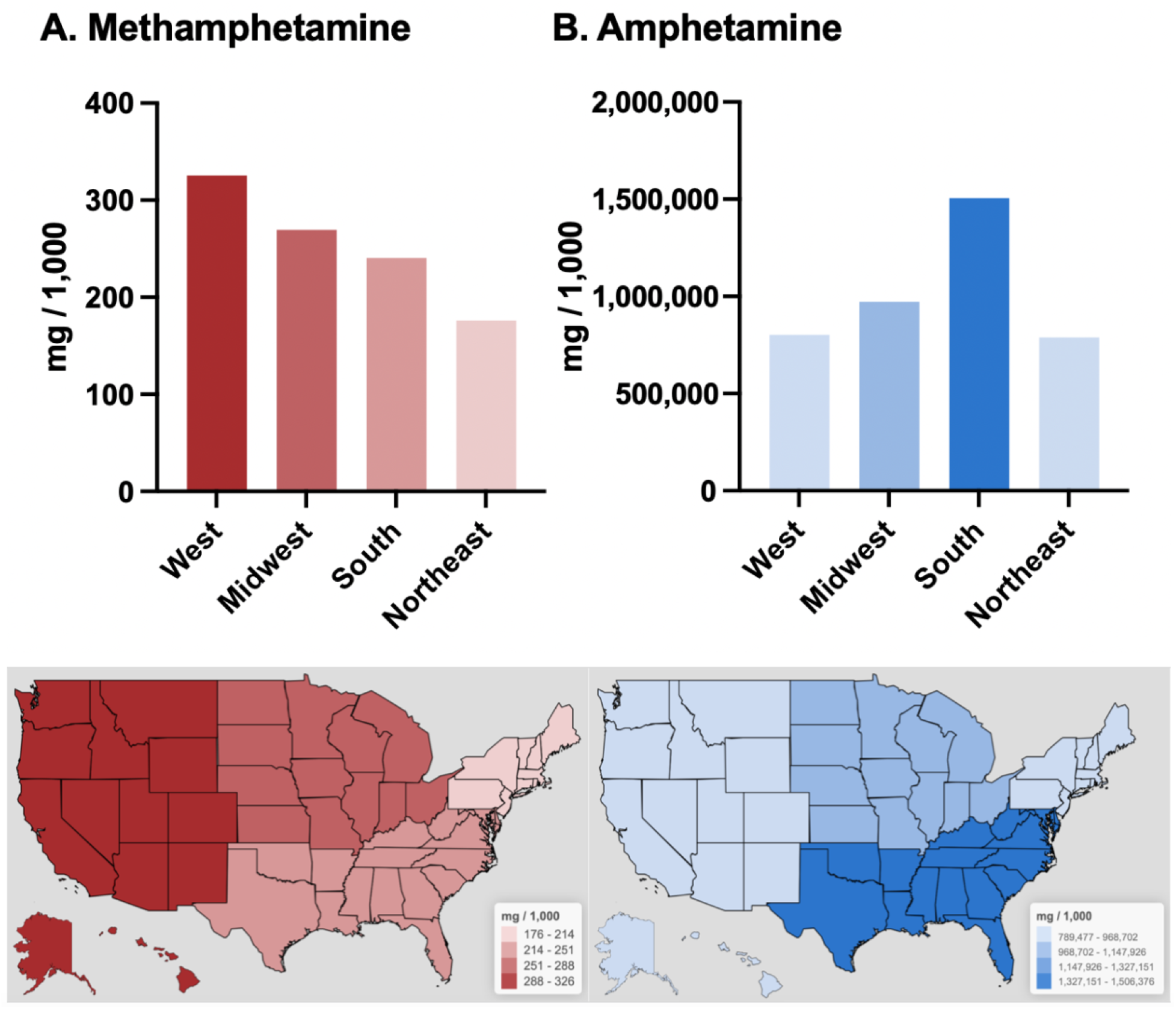
Total per capita drug weights of (A) methamphetamine and (B) amphetamine in milligrams per 1,000 people by region of the US in 2019. Heat maps of the total per capita drug weights of methamphetamine (bottom left) and amphetamine (bottom right) in milligrams per 1,000 people by region of the US in 2019.

For both methamphetamine and amphetamine, over 97% of retail drug purchases came from pharmacies. Of the remaining less than 3%, a majority of drug purchases for both substances were from hospitals. There were 39.1-fold more registrants that received amphetamine (66,475) than methamphetamine (1,702). The vast majority of both methamphetamine (97.30%) and amphetamine (97.60%) recipients were pharmacies. Hospitals accounted for 2.30% for methamphetamine and 2.37% for amphetamine recipients. Practitioners, mid-level practitioners, and teaching institutions only accounted for a combined total of 73 registrants, which made up the remaining 0.03% of amphetamine recipients. Comparing the total grams of each drug distributed via sales reported to ARCOS to the production quotas set for 2019, only 16.1% of methamphetamine allowed to be produced (for sale, not conversion) was distributed, while 54.0% of amphetamine allowed to be produced (for sale, not conversion) was distributed.

## 4. Discussion

There are 3 key findings from this novel pharmacoepidemiological report. The first being the regional variations in methamphetamine and amphetamine. The second being the four-thousand-fold difference between amphetamine and methamphetamine distribution, with amphetamine being most common. Finally, the third key finding is that for both schedule II substances as reported by ARCOS in 2019, distribution is remarkably lower compared to production quotas.

Methamphetamine distribution was found to be highest in the Western and Midwestern regions of the US. This was slightly different from regional amphetamine use trends of 2016, in which use in Western regions was one-third lower compared to Northeastern and Midwestern regions [14]. Our findings are consistent with previously found patterns of methamphetamine use that correlated with its location of illicit production from small toxic labs as well as superlabs [15,16]. However, despite the West and Midwest continuing to have the highest methamphetamine use rates, production and distribution of the drug is now believed to be more widespread across the US with doubling of illicit lab seizures in the South and fewer in the Northeast [16,17].

Rhode Island is one example of a state with one of the lowest distributions of methamphetamine, but one of the highest distributions of amphetamine, although this does not always follow the Northeast region’s patterns. It is consistent, however, with both the altitude-methamphetamine use correlation and another pattern. When *The Comprehensive Methamphetamine Control Act* (CMCA) of 1996 was enacted, which targeted illegal methamphetamine production but not pharmaceutical amphetamines, the use of prescribed amphetamines skyrocketed [18]. This pattern of low methamphetamine use with high amphetamine use was observed in a Rhode Island Hospital where MDMA usage was more common than methamphetamine usage, which was zero during the time of the study, among trauma patients, but the incidence of MDMA and amphetamine only was the same [19]. Based on the findings from this study, this inverse pattern of drug use or distribution is also applicable at the regional level. These results are also consistent with state psychostimulant (methylphenidate and amphetamine) distribution rates of 2000 in which Rhode Island oscillated within the higher quartile and 75th percentile above the national average [10].

The results from this study highlight variations in drug distribution across the United States, with one of the most recognizable disparities being that of Montana in which no methamphetamine drug distribution was reported in 2019. Patterns of increased drug use in the West and Midwest including states such as Washington, Oregon, Utah, Wyoming, Colorado, North Dakota and Minnesota, may be explained by increases in adult ADHD cases due to revision of ADHD criteria; in which DSM-5 symptom domain thresholds provide better identification of adult ADHD symptoms and associated impairments than DSM-IV, which is most effective in setting the criteria for young children [20]. Lower drug distribution by weight can be noted in the states in the Southern and Northeastern regions of the country, such as Texas, Arkansas, Tennessee, Vermont and West Virginia. These results differ appreciably from what has been previously reported in which US amphetamine distributional variations from 2010-2017 demonstrated that the overall +67.5% rise was less pronounced in the West compared to other regions [21]. Additionally, the lowest values of Daily Dosage/population (20 mg/day/person for amphetamine) were also reported within the Western region [21]. Previous studies using data from the National Health Interview Survey (NHIS) by the CDC found that from 1997-2016, weighted prevalence of ADHD diagnosis was highest in the Northeast (10.8-13.6%, 95 CI) and lowest in the West (6.1-7.8%, 95CI) [22]. This is consistent with other findings in which highest rates of ADHD diagnosis and medication use in children aged 3-17 years old for 2018-2019 were highest in Southern regions (10.97% and 7.52%, respectively) and lowest in Western regions (7.46% and 4.14%, respectively) [23].

Although Montana resides within the Western part of the United States and is surrounded by states showing highest rates of methamphetamine drug weight distribution per 1,000 people, there was zero distribution for 2019. This deviation in methamphetamine drug distribution may be explained by the stigmatization of methamphetamine use and amphetamine-based medications used to treat ADHD, which may be discouraging physicians from recommending them as a therapeutic option. Another plausible explanation for the regional variations in amphetamine and methamphetamine distribution across the US could correlate to regional concentration of younger physicians who are on average more likely to over-prescribe psychostimulants compared to older colleagues [10]. Furthermore, regional differences in medical specialties could also account for these variations as areas with larger populations of psychiatrists could account for increased psychostimulant distribution [10].

The Drug and Chemical Evaluation Section on Methamphetamine provided by the DEA indicates there is only one current licit methamphetamine product, Desoxyn, which is available in 5mg, 10mg, and 15mg tablets that is FDA-approved in managing ADHD and Obesity [24]. The FDA access data on Desoxyn (methamphetamine hydrochloride tablets) warms about limiting use for ADHD and Obese patients, and consideration of prescribing Desoxyn should come after other alternative pharmcotherapies, or treatment programs have been deemed unsuccessful [25]. The limited use of methamphetamine in treating ADHD is consistent with the prevalence of amphetamine distribution in this pharmacological report.

The increasing popularity of telemedicine may also provide insight upon methamphetamine and amphetamine distribution trends. These health platforms connect patients with healthcare professionals online allowing for increased access to care by removing physical barriers in seeking treatment. One retrospective cross-sectional study on encounters for ADHD via Direct-to-consumer telemedicine found that from 2016 to 2018, ADHD visits increased 500%, with 43% of ADHD related encounters resulting in prescription of non-controlled medications [26]. Similar telemedicine platforms such as Cerebral Inc. offer online prescriptions to controlled medications such as amphetamine-based stimulants and benzodiazepams, which has led to US federal investigation on prescribing practices [27]. Over-prescribing of these controlled medications by telemedicine companies may explain distributional variations. Additionally, a cross-sectional study conducting analyzing the accessibility of amphetamine-dextroamphetamine through digital pharmacies, it was determined that of the 62 pharmacies found via popular web search engines, 61 pharmacies were determined to be rogue and unclassified, 100% of which were selling drugs without a prescription, quantity limit, or requirement of a health-related assessment [28]. Illegitimate online pharmacies and telehealth companies overprescribing controlled substances with high potential for abuse can exacerbate diversion and nonmedical use of amphetamine and methamphetamine.

Stigmatization of drugs such as amphetamine and methamphetamine extends beyond the nature of their non-medical uses as it also encompasses the stigma around certain health conditions such as HIV and other STIs, including adult ADHD diagnoses. Studies regarding stigma around mental health disorders, has shown that the public tends to believe that ADHD symptoms reported are often autonomous, leading them to discredit and undermine the affected individuals diagnosis [29]. Adults suffering from ADHD are often labeled as infantile and socially objectionable, in this form, public stigma breeds self-stigma in those with ADHD and in order to avoid peer-rejection, sufferers prefer to conceal symptoms [29,30]. Additionally, this view of ADHD symptoms being a character flaw or weakness contributes to an overall negative self-image and encourages individuals to increase social withdrawal [30].

Stigmatization ultimately prevents ADHD sufferers from receiving treatment and allowing for mild symptoms to develop into severe psychiatric disabilities impacting social functioning [29,30]. In those who do seek medical treatment for ADHD, they are also susceptible to stigmatization and public mistrust of the medications available to them, specifically stimulant drugs such as methamphetamine. Among the most common concerns of ADHD medication is the risk of children and adults developing substance misuse disorders from exposure to stimulants. Stigmatizing attitudes in regard to ADHD medication make children and adults less likely to adhere to treatment and promotes noncompliance, which also negatively affects patients exhibiting help-seeking behaviors despite strong support and evidence that combined treatment of medication and psychotherapy is effective in ADHD management [29]. Even in cases in which substance misuse co-occurs with ADHD, stimulant drugs are still effective in treating ADHD and do not increase misuse potential for other drugs [31].

Individuals suffering from ADHD are at increased risk of also suffering from substance use disorders (SUD) with a prevalence of 10-24% in adults [31]. There exists the misperception that this occurs due to ADHD medication dependence rather than due to dopamine neurotransmission, a neurotransmitter also heavily implicated in addiction and associated neurobiological changes. Dopamine plays a major role in motivation and reward processing as well as reinforcing drug effects, leading to compulsive behaviors in drug users [32]. In considering the impact that ADHD has on SUD, issues regarding stimulant based pharmacotherapy are presented. A major fact that is considered is how detrimental untreated ADHD is in patients with SUD, as proper management can prevent social deficits, lack of educational and job attainment, and development of drug addictions. Individuals suffering from both ADHD and SUD are less likely to adhere to rehabilitation treatments despite increased exposure compared to those without ADHD [31]. Furthermore, substance misuse begins at a younger age in those with ADHD and remission is harder to achieve [31]. When both comorbidities are treated, there exists better outcomes. Although amphetamine-based medications have higher misuse potential in patients with SUD, substitution with sustained-release methamphetamine demonstrated reduced positive urine samples and cocaine cravings in a placebo-controlled randomized clinical trial [33].

Health-related stigmas, such as in SUD, lead to the rejection and humiliation of people suffering from diseases that have been labeled by socio-cultural standards as unrespectable health conditions. Additionally, stigmas also negatively impact services offered to patients to treat their conditions if they are associated with addictive drugs such as methamphetamine, amphetamine, and methadone [34]. Intervention methods geared towards reducing stigma around SUDs, prevent self-stigma and destructive behaviors by those devalued by the public. They aim to educate and build social acceptance by the public, and in turn prevent substance misuse more effectively than stigmatization does. In restructuring these attitudes about substance misuse, negative stereotypes guiding public policy, allocation of health-care expenses, and treatment of patients by physicians can be reduced [34].

Positive attitudes towards substance misuse treatment allows for prevention of infections and STIs commonly associated with risky behaviors and lack of hygiene in SUD via harm reduction intervention methods. Intravenous administration of drugs such as methamphetamine, account for 60% of emerging Hepatitis C cases and one-third of new HIV transmissions; providing those who are unable to stop using drugs with sterile syringes, allows for safer syringe practice [35]. Providing sterile syringes via exchange has been reported to be most effective in reducing HIV transmission. Furthermore, it allows for a foundation for other forms of intervention and treatments to be promoted such as methadone maintenance in treating heroin addiction [35].

In relation to the methamphetamine distribution data in Montana, which was zero, the effect of the Montana Meth Project (MMP) can serve as an explanation especially due to the fact that amphetamine data is still within the lower range when compared to other states. In comparing amphetamine and methamphetamine distribution within this region, we can speculate that lack of distribution may be from stigmatization of both prescription and illicit methamphetamine use caused by the MMP initiative. The objective of the MMP was to educate and discourage youth from using illicit meth by stigmatizing use and making it socially unacceptable. As previously discussed, this stigmatization, in combination with heavy advertising against amphetamine use through stereotyped images of what people on meth look like, may be preventing health care providers from recommending it as ADHD treatment due to fear of public push back [36]. Studies have shown that social stigma serves as a major effector in physicians not recommending certain drugs and patient refusal to take them [37]. Furthermore, results of a study conducted using data from 1999-2009 Youth Risk Behavior Surveillance System indicated that the project’s impact on curbing meth use was statistically non-significant [36].

## 5. Conclusion

This study evaluated the regional variations in methamphetamine and amphetamine drug distribution within the United States. Disparities in drug distribution data as extracted through the comprehensive ARCOS database demonstrated that methamphetamine in the Western and Midwestern portions of the country (excluding Montana) showed increased drug distribution when compared to other parts. In terms of amphetamine data, the opposite was true, in which distribution was lowest in the Western region. The most notable was the state of Montana in which methamphetamine distribution was zero. We also found that amphetamine distribution was four-thousand fold more common than methamphetamine. These variations in distribution are likely present due to stigmatization and accessibility differences to amphetamine-based medications. In addressing stigma revolving ADHD diagnosis and treatment options as well as assessing intervention options for comorbidities such as SUD, distribution may change as public health policies and health care provider recommendations are heavily correlated to public opinion. Future research must be done in order to quantify the effect that initiatives such as the Montana Meth Project have on health care provider recommendation of ADHD stimulant medications. Future studies may also aim to investigate possible causes for the disproportion between distribution and production quotes as reported by ARCOS in 2019. Furthermore, identifying unique prescribers of methamphetamine may also explain the regional variations and distribution trends present.

## Data Availability

Publicly available datasets were analyzed in this study. All data produced are available online at

https://www.deadiversion.usdoj.gov/arcos/retail_drug_summary/report_yr_2019.pdf

https://deadiversion.usdoj.gov/fed_regs/quotas/2018/fr1228.htm

## Author Contributions

Conceptualization, Data curation, Formal analysis, Funding acquisition, Investigation, Methodology, Project administration, Resources, Software, Supervision, Validation, Visualization, Writing - original draft, Writing - review & editing. All authors have read and agreed to the published version of the manuscript.

## Institutional Review Board Statement

Procedures were exempt as reviewed by the IRB of Geisinger.

## Informed Consent Statement

Not applicable.

## Data Availability Statement

Publicly available datasets were analyzed in this study. This data can be found here: https://www.deadiversion.usdoj.gov/arcos/retail_drug_summary/report_yr_2019.pdf

https://deadiversion.usdoj.gov/fed_regs/quotas/2018/fr1228.htm

## Supplemental Tables

**Supplemental Table 1.**
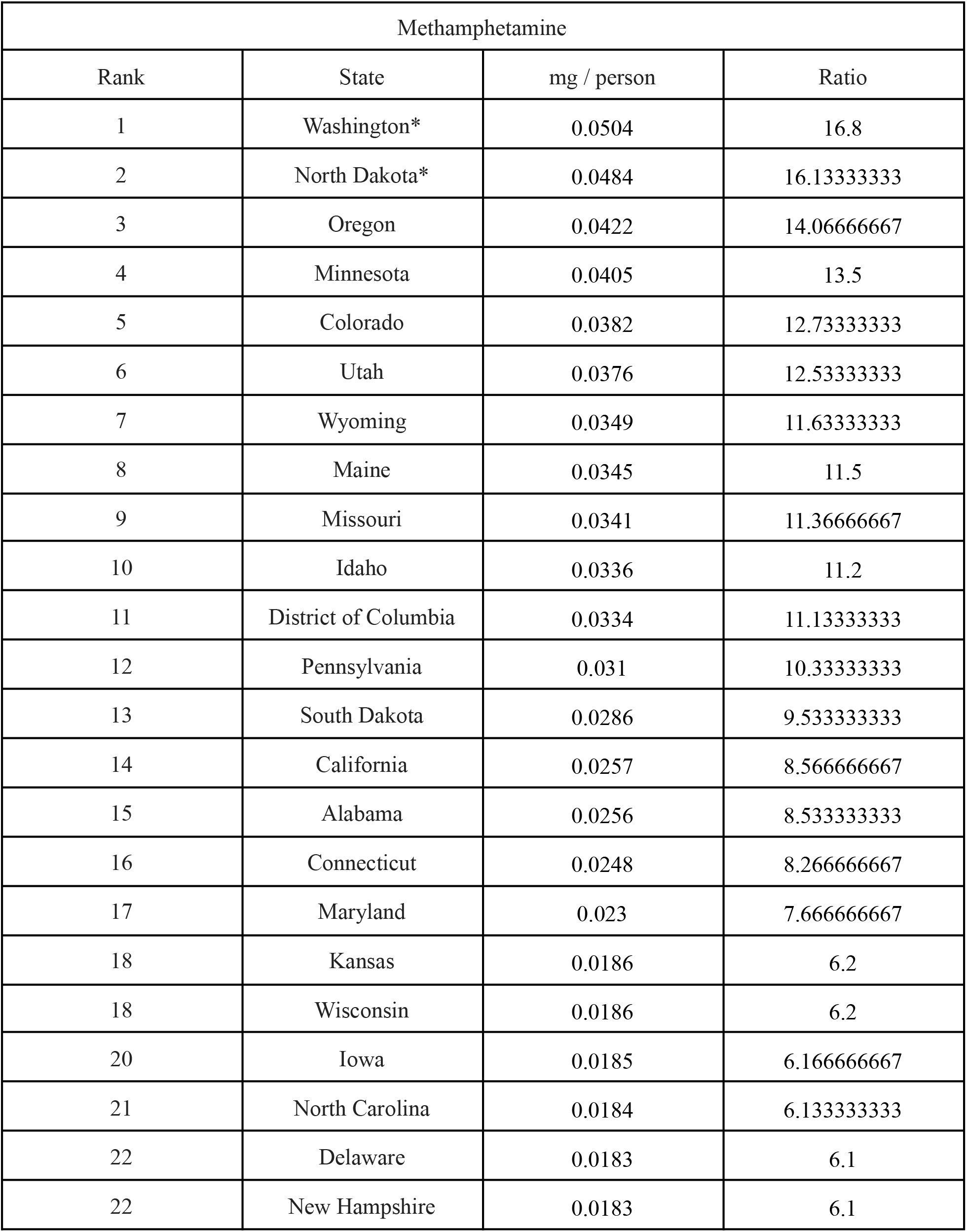
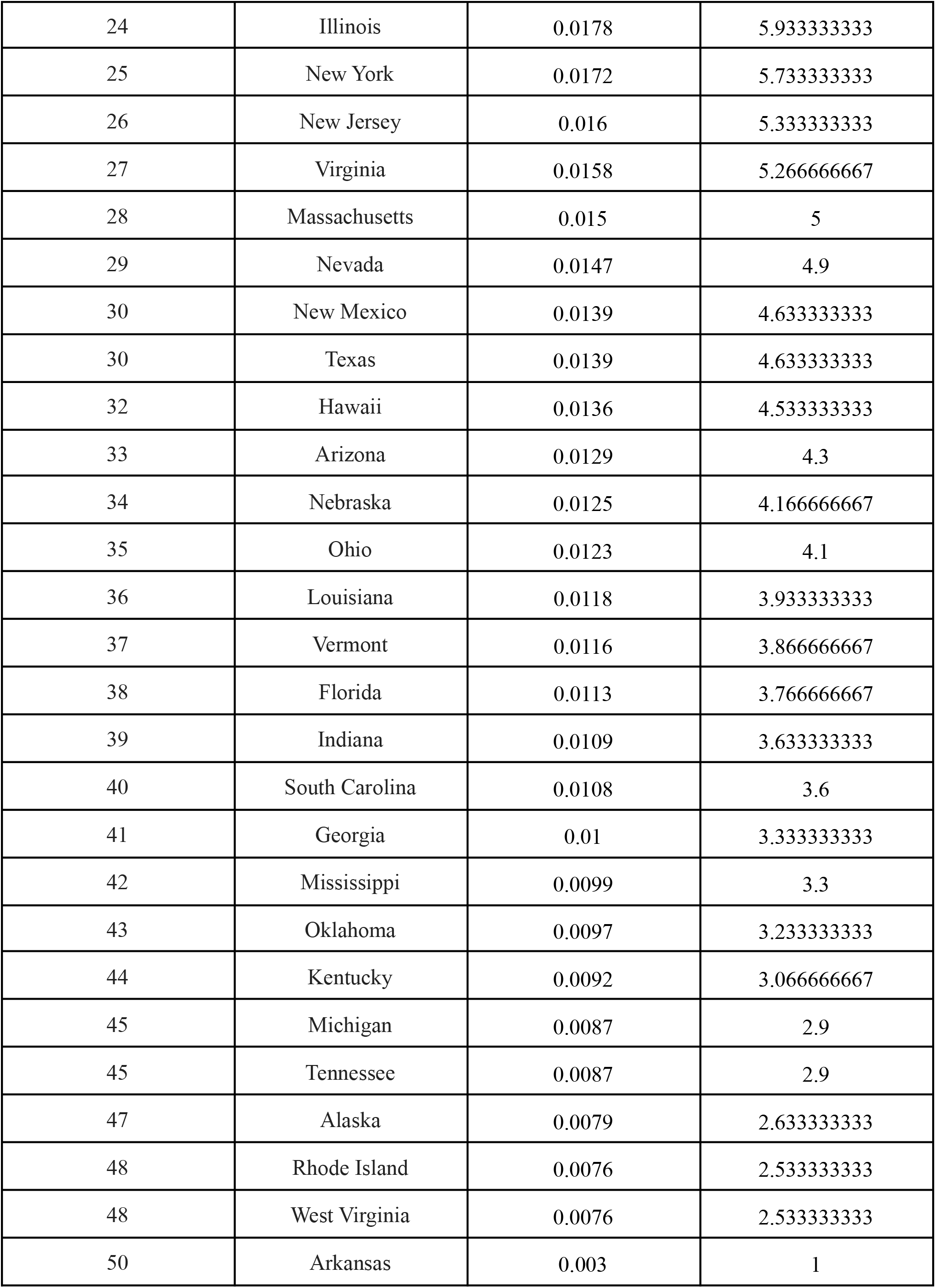
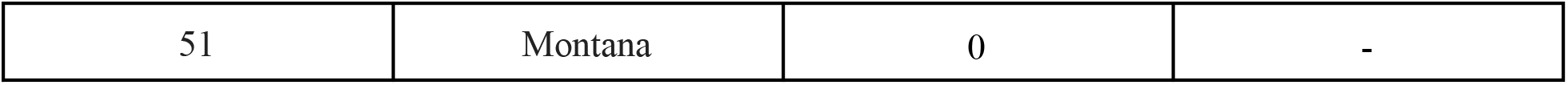
Per capita distribution in mg/person for all 50 states and Washington DC, and per capita distribution, expressed as a ratio relative to the lowest, nonzero state, ranked from highest to lowest for methamphetamine. Montana ranked 51st, but was not used as the state with the lowest per capita distribution as its value was zero. *States with significant values outside the 95% Confidence Interval (Mean = 0.01983, SD = 0.01197).

**Supplemental Table 2.** Per capita distribution in mg/person for all 50 states and Washington DC, and per capita distribution, expressed as a ratio relative to the lowest, nonzero state, ranked from highest to lowest for amphetamine. *States with significant values outside the 95% Confidence Interval (Mean = 79.7947, SD = 27.7818).

| Amphetamine |  |  |  |
| --- | --- | --- | --- |
| Rank | State | mg / person | Ratio |
| 1 | Louisiana* | 153.76 | 4.918132415 |
| 2 | Rhode Island* | 150.0954 | 4.800917352 |
| 3 | Utah* | 139.1981 | 4.452358791 |
| 4 | New Hampshire | 131.1295 | 4.194278385 |
| 5 | South Carolina | 126.4673 | 4.045154315 |
| 6 | Massachusetts | 116.013 | 3.710765452 |
| 7 | Alabama | 114.873 | 3.674301671 |
| 8 | Michigan | 102.2996 | 3.272131756 |
| 9 | Missouri | 101.6732 | 3.252095868 |
| 10 | North Carolina | 97.8915 | 3.131135271 |
| 11 | Delaware | 96.976 | 3.101852296 |
| 12 | Virginia | 95.1194 | 3.04246751 |
| 13 | Kansas | 93.9553 | 3.005232872 |
| 14 | Wisconsin | 89.4636 | 2.861562377 |
| 15 | Indiana | 88.6052 | 2.83410579 |
| 16 | Tennessee | 88.4453 | 2.828991265 |
| 17 | Minnesota | 87.746 | 2.806623614 |
| 18 | District of Columbia | 85.9268 | 2.748435096 |
| 19 | Mississippi | 85.3482 | 2.729928128 |
| 20 | Georgia | 85.1263 | 2.722830485 |
| 21 | Maine | 79.051 | 2.52850732 |
| 22 | Idaho | 77.7875 | 2.488093296 |
| 23 | Texas | 76.3097 | 2.440824721 |
| 24 | Arkansas | 75.7318 | 2.422340143 |
| 25 | Colorado | 75.3007 | 2.408551076 |
| 26 | Ohio | 74.4264 | 2.380585915 |
| 27 | South Dakota | 73.3583 | 2.346421912 |
| 28 | Washington | 72.933 | 2.332818362 |
| 29 | Florida | 72.4939 | 2.31877341 |
| 30 | Iowa | 71.9545 | 2.301520284 |
| 31 | Oregon | 70.6466 | 2.259686092 |
| 32 | Maryland | 70.5241 | 2.255767834 |
| 33 | Pennsylvania | 70.4767 | 2.254251709 |
| 34 | Kentucky | 69.1394 | 2.211477135 |
| 35 | Vermont | 67.5655 | 2.161134727 |
| 36 | Connecticut | 67.1301 | 2.147208122 |
| 37 | Illinois | 65.8648 | 2.106736524 |
| 38 | Montana | 64.907 | 2.07610055 |
| 39 | Nebraska | 64.7484 | 2.071027607 |
| 40 | Oklahoma | 60.3031 | 1.928841251 |
| 41 | Arizona | 60.2187 | 1.926141652 |
| 42 | North Dakota | 59.1006 | 1.89037836 |
| 43 | New York | 54.8047 | 1.752970679 |
| 44 | New Jersey | 53.2118 | 1.702020541 |
| 45 | West Virginia | 51.9401 | 1.661344234 |
| 46 | Wyoming | 51.5251 | 1.648070138 |
| 47 | Alaska | 48.7033 | 1.557812685 |
| 48 | Nevada | 42.3491 | 1.354568688 |
| 49 | California | 33.9899 | 1.087193216 |
| 50 | Hawaii | 31.6573 | 1.012583203 |
| 51 | New Mexico | 31.2639 | 1 |

